# Social-media based Health Education plus Exercise Programme (SHEEP) to improve muscle function among young-old adults with possible sarcopenia in the community: a study protocol for intervention development

**DOI:** 10.1101/2023.05.18.23290177

**Authors:** Ya Shi, Emma Stanmore, Lisa McGarrigle, Chris Todd

## Abstract

**Introduction:** Prevalence of possible sarcopenia is estimated to be significantly higher in community-dwelling older adults than that of confirmed or severe sarcopenia. However, there are currently far fewer non-pharmacological intervention strategies for possible sarcopenia than for sarcopenia in the community. Meanwhile, one type of non-pharmacological intervention in sarcopenic area, health education, is under-researched, and older people’s awareness about sarcopenia is extremely low, necessitating an immediate dissemination tool for prevention. Social media may be a potential, scalable, low-cost tool for this. This study protocol outlines how a social media-based multicomponent intervention will be co-designed with stakeholders to address this evidence gap.

**Methods and analysis:** Guided by the Medical Research Council’s framework, the proposed research covers two phases that employ a co-design approach to develop a theory-based multicomponent intervention to increase sarcopenia prevention in the community. The participants will be recruited from young-old adults (60∼69) with possible sarcopenia in the community of Changsha, China. Maximum sample size will be 45 participants in total, with 18∼25 participants in the development phase and 15∼20 participants in the pre-testing phase. During two rounds of focus groups with older adults, a social-media based intervention strategy will be developed from a theory-based conceptual model and an initial intervention plan formulated by the research group. After this, there will be a three-week pre-testing phase, followed by a semi-structured interview to further modify the theory-based conceptual model and the social-media based intervention strategy. The focus of the data analysis will be on thematic analysis of qualitative data primarily derived from the group interview and the semi-structured interview with key stakeholders.

**Ethics and dissemination:** This study has been approved by the University of Manchester Research Ethics Committee (Project ID: 15664), and permissions have already been granted by collaborators in relevant Chinese organisations. We will collaborate with stakeholders to inform our dissemination strategy and co-present our findings (e.g., at community events or through social media). Furthermore, we will disseminate our findings to academics and healthcare professionals via webinars, academic conferences, and peer-reviewed publications.

**Strengths and limitations of this study:**

- This is the first study to develop a health education and exercise intervention aimed at improving muscle function in community-dwelling young-old adults with possible sarcopenia.
- This is the first study in the sarcopenic area to develop a multicomponent intervention for sarcopenia prevention based on social media (TikTok).
- This study addresses some shortcomings of single text or audio materials or lengthy lessons in current health education for sarcopenia with short videos.
- This study employs co-design to ensure that the intervention strategy is acceptable to relevant stakeholders and meets their needs by soliciting their input.
- The sample size will be small that it may not be representative of the entire Chinese older population.
- This study will not draw any conclusions about the intervention effectiveness due to the short duration of the pre-testing phase.
- The study findings are unique to the community setting and population in China and may not be generalised to other settings or countries.

## INTRODUCTION

Possible or probable sarcopenia is a relatively new classification proposed by the Asian Working Group for Sarcopenia (AWGS) 2019 and the European Working Group on Sarcopenia in Older People (EWGSOP2), which refers to low muscle strength^1,2^. The prevalence of possible sarcopenia is quite high among community-dwelling older adults. According to a latest population-based longitudinal study conducted in China, the estimated prevalence of older adults aged 60 and over with possible sarcopenia in the community was 46.0%^3^. Pérez-Sousa et al.^4^investegated 5237 Colombian older adults aged≥60 years and found that the prevalence of probable sarcopenia was as high as 46.5%. Several recent cross-sectional studies also reported a high prevalence of possible sarcopenia in older population living in the community, with 26.9% in Swiss (>60y)^5^, 25.4% in Greek (≥75y)^6^, 23.7% in Korean (≥65y)^7^. Besides, Wu et al.^8^ discovered that the prevalence of possible sarcopenia (38.5%) among older people in the community was significantly higher than that of confirmed (18.6%) and severe (8.0%) sarcopenia.

The young-old age group has a high prevalence of possible sarcopenia, but relatively fewer intervention methods, requiring additional focus. Up to 71% of the participants in the longitudinal study conducted in China who reported possible sarcopenia were between the ages of 60 and 70^3^. More than half (52%) of the older Colombians diagnosed with possible sarcopenia in the cross-sectional study were aged 60 to 69^4^. Wu et al.^9^examined that the prevalence of possible sarcopenia among adults aged 55 and older at 11 rural community daycare centres in Taiwan was as high as 68.7%. In addition, our previous scoping review of the non-pharmacological interventions for community-dwelling older adults with possible sarcopenia or sarcopenia revealed that 70-79-year-olds (64.8%) have been studied more than 60-69-year-olds (18.5%) between 2010-2023.03, and the number of interventions for possible sarcopenia (11.9%) was significantly lower than for sarcopenia (72.9%)^10^. However, both the EWGSOP and AWGS recommend possible sarcopenia as an important threshold to trigger assessment of causes and initiate intervention in medical practice^1,2^. Despite the fact that the definition of an older person’s age is not uniform around the world due to different conditions in different countries (e.g. ≥ 50 in Africa, ≥ 60 in United Nations and China, ≥65 in western countries, ≥75 in Japan), the ages of 60 and 65 are frequently used^11-13^. Moreover, different scholars interpreted the definition of “young-old” differently, such as 60-69 or 65-74^14,15^. Hence, based on the high prevalence of possible sarcopenia in the 60-69 age group, our scoping review’s identification of a deficiency in interventions for the 60-69 age group, and the fact that this study will be conducted in China, 60-69 is considered young-old for the purposes of this study.

Non-pharmacological interventions are essencial for sarcopenia prevention, but health education is under-researched. EWGSOP, AWGS, and some researchers recommend facilitating timely lifestyle interventions and related health education for primary health care in community and prevention settings, which will increase awareness of sarcopenia prevention and intervention in diverse health care settings^,1,2,16-18^. Nevertheless, our previous scoping review found that the proportion of intervention types containing health education component(s) to prevent possible sarcopenia or sarcopenia (15.5%) was significantly lower than other types, such as exercise (52.8%) or nutrition (34.5%)^10^. Most groups of health education (81.8%) provided education materials unrelated to sarcopenia, whereas only four groups mentioned health education content containing sarcopenic knowledge, which may be the primary resaon for unsatisfactory results of existing health education^10^. Furthermore, there were only three traditional forms of health education identified in this scoping review, including group-based classes, face-to-face interactions and leaflets/materials^10^, which appears to be significantly fewer than the forms of health education in other chronic diseases, like digital health education on diabetes management^19^ and social media as an educational platform on hypertension^20-22^. An integrative review indicated that technologies promoting health education for older adults in the community also included software, videos, mock-up and so on^23^.

In the context of an Internet era and in light of the increased use of digital tools during the covid-19 pandemic, social media may be a promising medium for health education to disseminate knowledge and raise awareness of disease prevention among older adults. A cross-sectional study examined the current knowledge of middle-aged and older adults regarding sarcopenia and showed that only 9% of participants (57.0∼75.1y) reported knowing what sarcopenia is, although 76% were willing to begin treatment in the event of a sarcopenia diagnosis and 71% were willing to prevent sarcopenia^24^. Therefore, effective strategies to rapidly increase sarcopenia awareness among community-dwelling older adults are urgently required. In the midst of the Covid-19 pandemic, people are living in an era of global communication in a hyper-connected society in which the transmission channels between individuals have been drastically altered by the internet and social networks in particular^25^.

Social media has changed our methods of communication, information exchange, and even impressions, and could be the fastest way to transfer knowledge of preventive practises and disseminate health or scientific information to healthcare professionals and others who require it^26-28^. Some types of social media have been proven to be effective in medical studies to disseminate health knowledge or improve behaviour, such as Twitter, Facebook, Instagram, TikTok, WeChat, Weibo^29-34^. More importantly, a number of studies have demonstrated that older populations are receptive to social media as a tool for knowledge dissemination or health intervention^35-28^. Particularly, TikTok, a platform for sharing short videos, has been phenomenal since the onset of the COVID-19 pandemic and has reached Chinese seniors^39^ by providing simple video-editing tools40and catering to specific informational and practical needs^41,42^. Hence, TikTok may be a valuable platform for attempting to develop a short-video-based intervention strategy to disseminate sarcopenia-prevention health education in older adults.

Apart from health education, this research programme was founded on behaviour change theories. Previous researches have already demonstrated that exercise could improve sarcopenic indices in sarcopenia patients^43-45^, but the maintenance of improvements in long-term behaviour change in community-dwelling older adults with sarcopenia is unclear10. Strong evidence indicates that successful behaviour change interventions in physical activity include several key components, such as theoretical framework, useful intervention components, and behaviour change techniques^46-49^. Behaviour change theories specify how a behaviour change intervention should function and guide intervention development, which can attempt to explain and predict behaviour change, generalise findings from prior work into novel areas, and refine and develop the theories themselves^,47,50-52^. Therefore, this study constructed a new conceptual model primarily guided by Michie et al’s Behaviour Change Wheel (BCW)^46^and the conceptual model for Lifestyle-integrated Functional Exercise using smartphones and smartwatches (eLiFE)^47^, with the objective of developing intervention content and monitoring process. BCW is a framework (Figure 1) mainly for developing intervention content for behaviour change, and several studies have demonstrated its efficacy and recommended its use in interventions for older adults^53,54^. In addition, the intervention phase was derived from the eLiFE conceptual model (Figure 2), which assists older adults in forming long-term physical activity habits through the use of a mobile app^47^, and the eLiFE programme was shown to be feasible and safe for young seniors^55^. We integrated the BCW and eLiFE conceptual models to create a new SHEEP conceptual model for preventing sarcopenia among older people (Figure 3).

**Figure 1.**
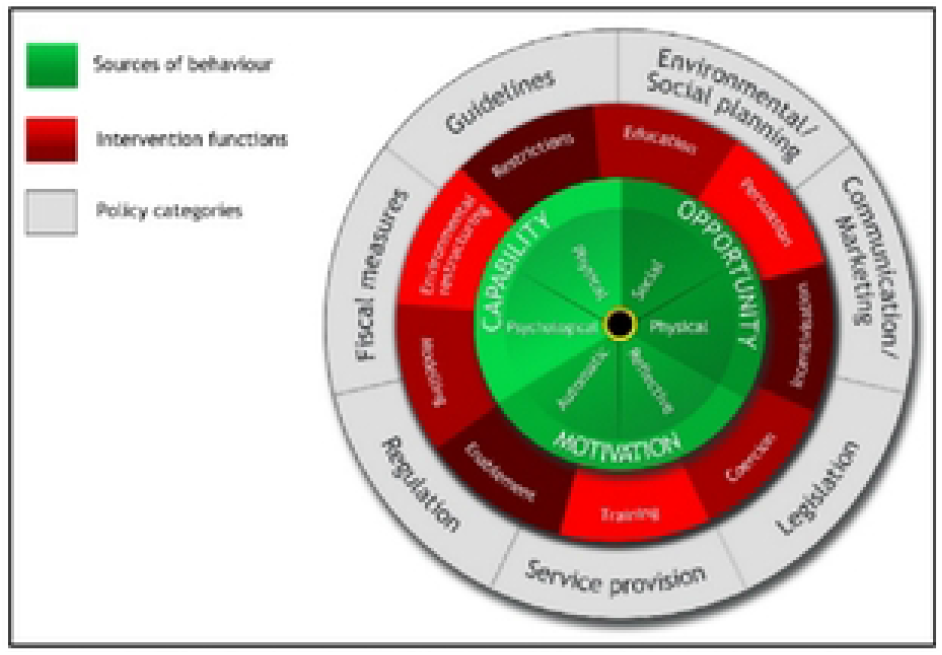
The Behaviour Change Wheel, Michie et al. 2011^46^

**Figure 2.**
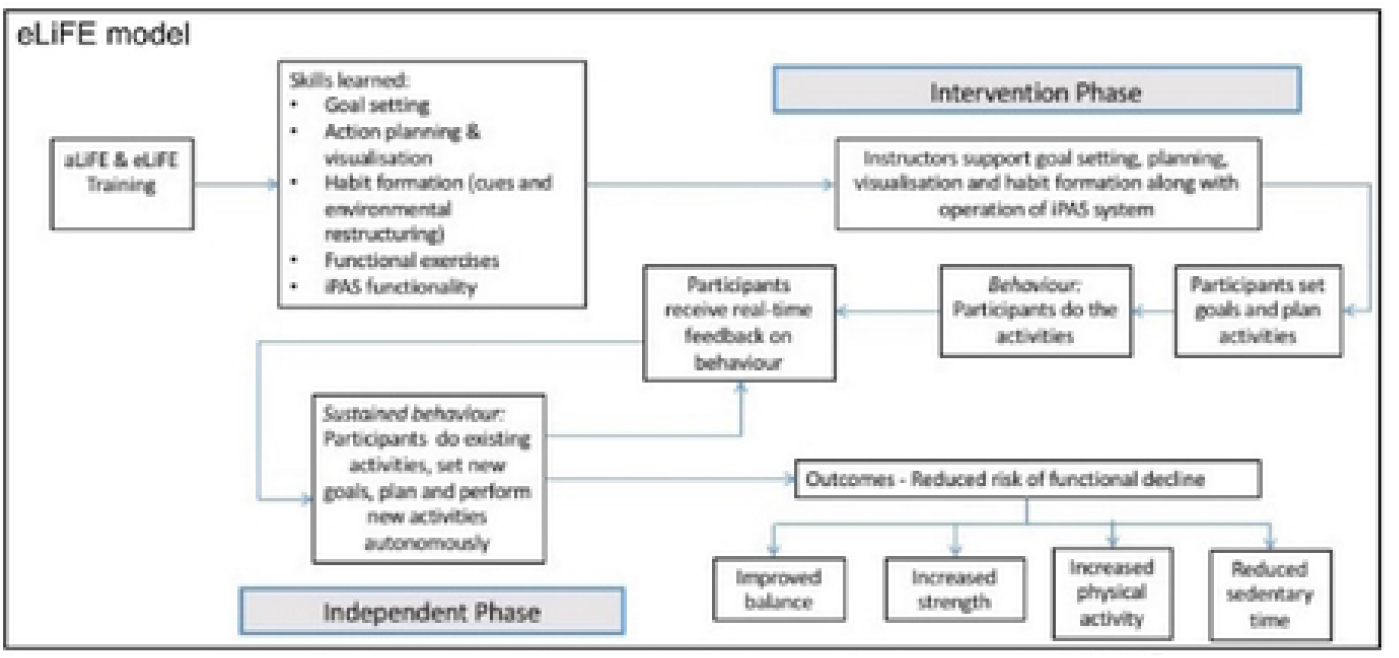
The eliFE Conceptual Model, Boullon el al. 2019^49^

**Figure 3.**
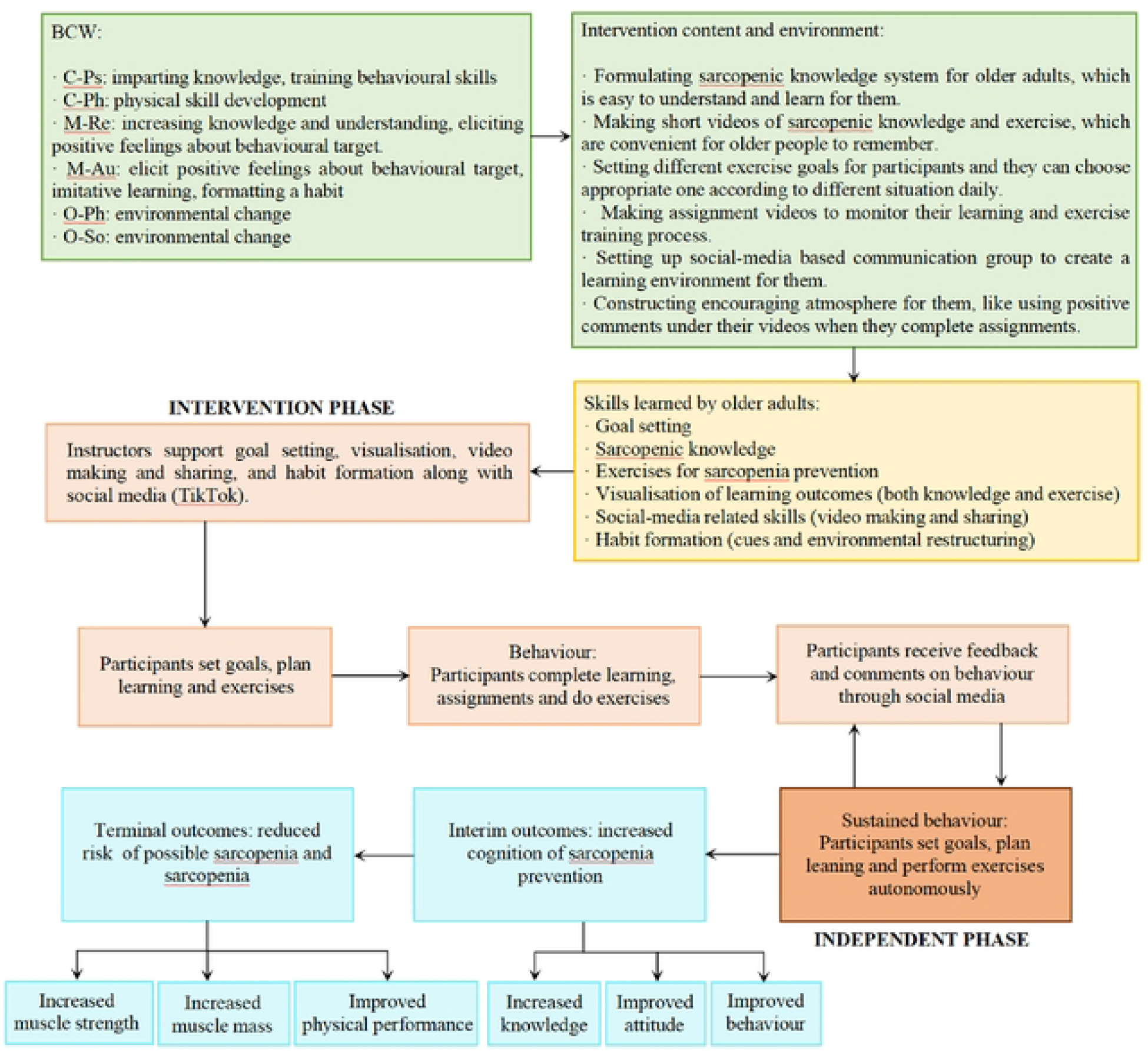
The SHEEP Conceptual Model. Note: C-Ps, psychological capability; C-Ph, physical capability; M-Re, reflective motivation; M-Au, automatic motivation; O-Ph, physical opportunity; O-So, social opportunity.

To improve the acceptability of the theory-based intervention strategy among older individuals, co-design method will be employed in this study. Strong evidence found that patients (even those close to death) and their families were consistently willing to engage in research^56^. Co-design in health care services typically refers to active collaboration between researchers, specialists, health care professionals, and non-academic partners such as patients or family members who are regarded as ‘experts of their experiences’^57-59^. In recent years, co-design methods have been utilised extensively in health care interventions, for example, a co-designed mHealth programme to promote healthy lifestyles^60^, a co-designed intervention to improve communication about the heart failure trajectory^61^, and co-designing complex interventions with people living with dementia and their carers^62^. Whether it is used for research or service improvement, co-design has been proved to be beneficial for study projects, users and services^,57,60-65^.

Overall, this research presents a protocol for the development of a theory-based and social-media based multicomponent intervention (health education plus exercise) by employing the co-design method for preventing sarcopenia in community-dwelling young-older adults. The aims of the research are: 1) To co-design an intervention strategy based on the SHEEP Conceptual Model; 2) To conduct a pre-testing stage to refine both the intervention strategy and the SHEEP Conceptual Model. In order to accomplish the research objectives, the following research questions will be posed: 1) What intervention contents should be included in different part of health education and exercise training? 2) How to ensure the correct dose of health education and exercise (in terms of frequency, intensity, and duration) can be achieved using a social-media based approach?

## METHODS AND ANALYSIS

### Study design

The SHEEP programme was structured under the guidance of the UK Medical Research Council (MRC) for developing and evaluating complex interventions^66^. This research protocol outlined the initial stage (intervention development) in the MRC’s guidance. Co-design method is the most crucial technique for completing the formulation of a social-media based intervention strategy to prevent sarcopenia among targeted population in this project. According to the five guiding principles and systematic framework proposed by Leask et al.^67^ for designing, implementing, and evaluating co-designed public health interventions, consideration is given to first completing the designing phase in this study, in order to formulate a social-media based intervention strategy that incorporates the perspectives of the relevant stakeholders. This includes the preparing, developing, and pre-testing phases (Figure 4).

**Figure 4.**
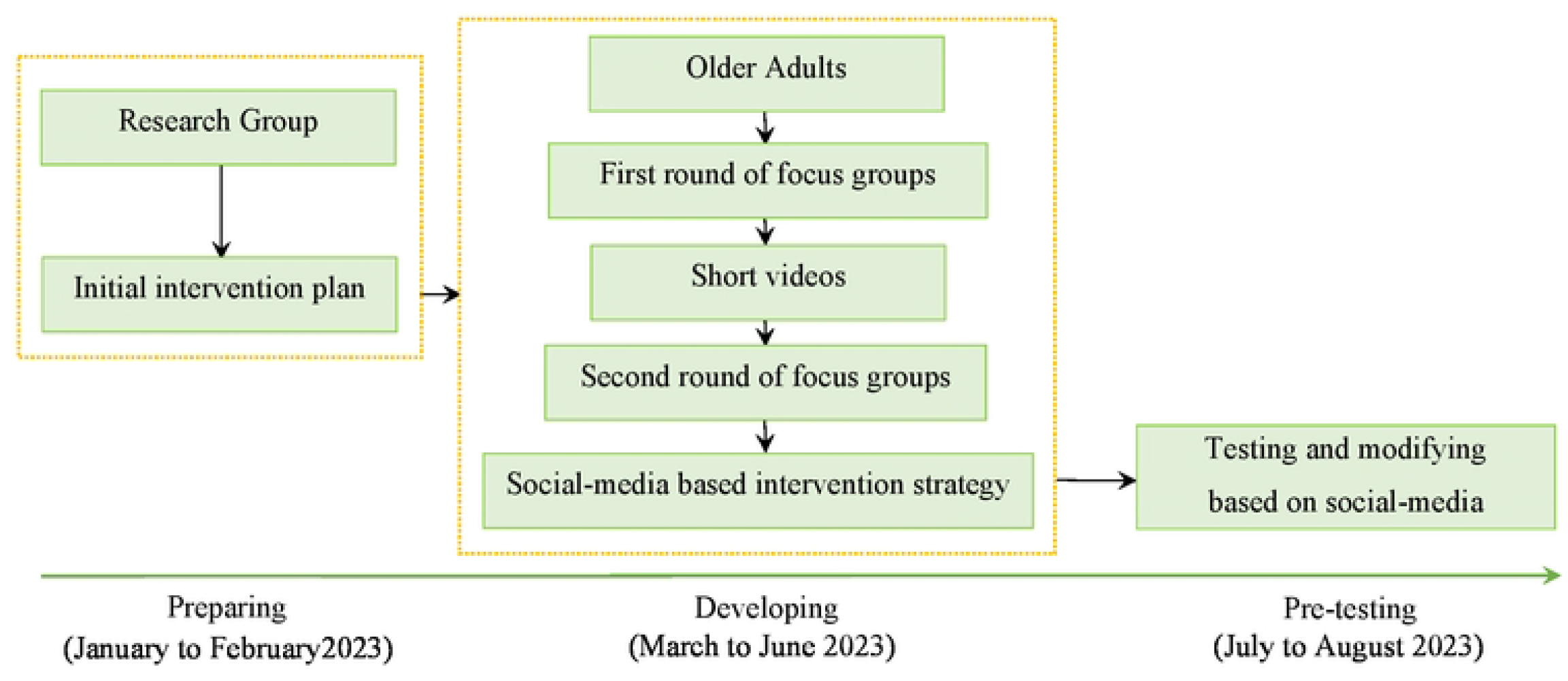
The Flowchart of This Research Protocol

#### Preparing

In January and February of 2023, the research team will develop an initial intervention plan based on literature reviews, the SHEEP Conceptual Model, and consultations with specialists regarding health education and exercise contents, frequency, intensity, and duration.

#### Developing

Between March and June of 2023, there will be two rounds of focus groups in the development procedure. Each round will include three focus groups, to which older adults with possible sarcopenia will be invited. The aim of the first round of focus groups is to discuss the initial intervention plan’s flaws with participants and adapt it to the needs of the target population; we will then create short videos based on the revised plan. The purpose of the second round of focus groups is to modify the content of each short video further.

#### Pre-testing

During July and August of 2023, a pre-testing process will be conducted with the goal of refining the SHEEP conceptual model and optimising the social-media-based intervention strategy and intervention process.

### Research setting

The entire study will be conducted at the Community Nursing Department of Xiang Ya Nursing School, Central South University, or within their cooperative community in Changsha, Human Province, China. It should be noted that the recruitment process will take place in a location that participants prefer based on their individual needs, such as the home of older participants or an office provided by our collaborator at Xiang Ya Nursing School or in the community. In addition, the co-design process will take place in an office provided by Xiang Ya Nursing School or the community, depending on the preference of the majority of participants within each focus group. However, if face-to-face focus groups are prohibited by local policy due to an outbreak, we will switch to online focus groups using Tencent Conference Software.

#### Research Participants

Participants must satisfy the following inclusion criteria: (1) 60∼69 years old; (2) Chinese residing in the community; (3) using TikTok prior to recruitment; (4) individuals with possible sarcopenia, as defined by low Grip Strength [M:<28 kg, F:<18kg]^2^, which will be measured with a digital handheld dynamometer (EH101, Xiangshan Inc, Guangdong, China); (5) informed consent to screening and research. The following are the criteria for exclusion: (1) unable to communicate or independently complete learning on TikTok in Chinese; (2) with serious or unstable medical illness, such as severe cardiovascular or respiratory conditions, mental disorder, dementia, etc.; (3) sufficient physical activity (≥ 150mins/week)^68,69^.

#### Sampling method and sample size

Purposive sampling will be used to select relevant stakeholders. To avoid bias against subgroups, we will select older adults based on their gender (male and female) and age (60∼65 and 66∼69). We aim to recruit a maximum sample of 45 older adults in total. For developing phase, we will recruit 18∼25 older adults to participate in two rounds of focus groups (with a 10%∼15% attrition rate). Each round will consist of three focus groups with a minimum of six participants in each focus group, based on the recommendations of Leask et al^67^ that 10∼12 co-creators in total is advised in co-design research and the Guidelines for Conducting a Focus Group that suggests six to ten people will be appropriate for a focus group70. Besides, we require 15∼20 additional participants for the pre-testing phase. In the two phases of our study, participants have the option of participating in only focus groups, only pre-testing, or both focus groups and pre-testing.

#### Recruitment, consent and withdrawal

With the assistance of the cooperative community health centre’s medical staff, we will first employ a population-based method to identify suitable subjects. First, we will visit a community health centre and inform health professionals about the case finding method, in which older individuals with the following clinical conditions are susceptible to possible sarcopenia: functional decline or limitation; unintentional weight loss; depressive mood; repeated falls; malnutrition; chronic conditions like heart failure, chronic obstructive pulmonary disease, diabetes mellitus, chronic kidney disease, etc. Second, we will only request that health professionals identify potential participants, not that they provide us with access to their medical records. We will provide health professionals with recruitment leaflets, and they will assist us in distributing the leaflets to potential population who visit the community health centre. Third, we will also send electronic recruitment leaflets to community WeChat groups through the assistance of health professionals. If potential participants are interested in this study after reviewing the leaflet, they can contact the researcher via the email address or telephone number listed on the leaflet. Researchers will meet with older adults in their homes or at the community health centre, based on their preference, and then present our research in depth in person. If they are truly interested, we will implement a screening assessment to ensure they meet the aforementioned eligibility requirements.

Potential participants who meet the eligibility requirements will be provided with a Full Explanation of Research that informs them of the detailed research content, the confidentiality of personal information, the potential benefits, the potential risks, and a coping strategy for the risks, should they wish to know. A Participant Information Sheet will be provided to subjects who are still interested and willing to participate in the study, and time will be allowed for consideration (at least 24 hours). The study will only continue with participants who provide informed consent after receiving all pertinent information.

As for the principle of withdrawal, participants will be permitted to withdraw at any time without providing a reason, and a withdrawal case report form (CRF) will be filled out to document the date and reason (if provided) for withdrawal. If participants have already participated in the focus group or in the pre-testing phase, their contribution will not be retracted as it contributes to a broader discussion; however, no excerpts from their contributions during the discussions will be used in any report or presentation. Besides, researchers will evaluate participants’ capacity to participate in the study and complete the intervention; if they are deemed incapable of providing consent, they will be withdrawn from the study, but the data already collected will be retained.

#### Study procedures

We will adhere to the five key principles and systematic framework for co-designed public health interventions proposed by Leask et al.^67^ throughout the entire research procedure. The five key principles are defining the purpose of the study, sampling, manifesting ownership, defining the procedure, and evaluating (the process and the intervention), all of which were structured into multiple sections to provide a systematic framework for the iterative co-creation process, including planning, conducting, reflecting, evaluating, and reporting (Figure 5). As the study’s sole objective is intervention development, we have modified our study procedure into three phases: pre-paring, developing, and pre-testing, while still adhering to the aforementioned rules of principles and systematic framework.

**Figure 5.**
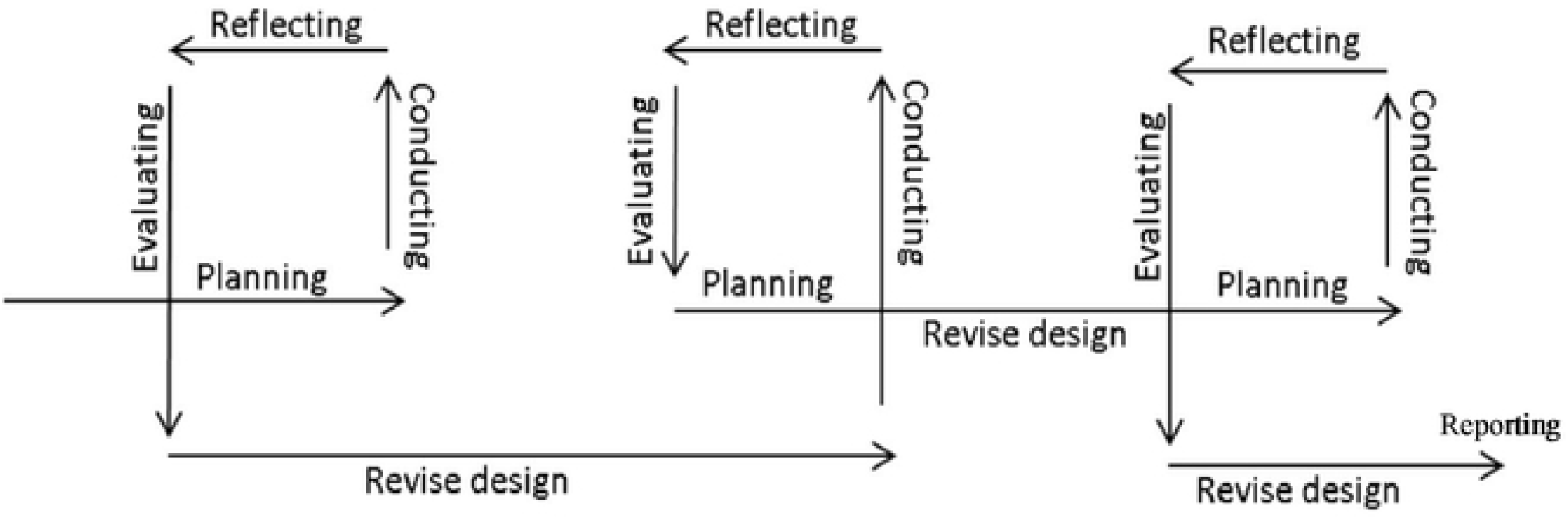
The Iterative Co-creation Process, modified from Leask ct al 2019^67^

#### Pre-paring phase (initial intervention content, frequency and duration)

Based on previous literatures and the SHEEP conceptual model, we have formulated an initial intervention plan that includes health education and exercise, which will be modified in January and February 2023 after consultation with specialists in the field of sarcopenia and exercise for older adults.

Regarding health education, the curriculum would cover nine topics: What exactly is sarcopenia? What is sarcopenia’s prevalence in older adults? What are the negative effects of sarcopenia? What factors contribute to sarcopenia? What clinical symptoms are associated with sarcopenia? What should you do if you suspect sarcopenia? What effects does physical activity have? What physical activities can help prevent sarcopenia in older adults? What other evidence exists for preventing sarcopenia in older adults living in the community? The contents will be modified and expanded in the future, and will also be presented in short videos with language that Chinese older adults can easily comprehend, following the development of focus groups. Currently, the health education frequency is set to 3 times/week and the duration per video is set to less than 5mins; however, these parameters will be modified after the developing and pre-testing phases.

In terms of exercise, resistance exercise (RE) will be the primary form of training for sarcopenia prevention, but specific motions must be selected and adapted in the co-design stage based on the characteristics of the Chinese older population. Systematic review and meta-analysis provided robust evidence that RE programmes can improve muscle mass and strength in middle-aged and older adults^71,72^. Aside from RE, we also consider a combination of other exercise modes [e.g. balance training (BT) and aerobic training (AT)], as older adults with sarcopenia are also likely to be at an increased risk for falls^73^ and exhibit reduced cardiorespiratory fitness^74^. One study proposed a pragmatic person-centred approach to guide individualised RE prescription for older adults with sarcopenia^75^, which will assist us in customising RE for Chinese older adults in this study, as illustrated in Table 1. Frequency and duration of exercise are progressive from the beginning to the end of the intervention, with frequency increasing from 3 days/week to 4 days/week and duration increasing from 20mins/session to 40mins/session, as the National Health Service (NHS) and Centres for Disease Control and Prevention (CDC) recommend older individuals at least 2 days/week and 150mins/week of activities to strengthen muscles^68,69^. Similarly with health education, all exercise components will be refined and produced as short videos following the developing phase.

**Table 1.** RE prescription for older adults with sarcopenia, Hurst et al 2022^75^

| Dimensions | Criteria |
| --- | --- |
| Principles of exercise training | specificity, overload and progression |
| Training frequency | two sessions per week |
| Exercise selection | lower body (Squat/leg press, Knee extension, leg curl, calf raise), upper body (chest press, seated row, pull down) |
| Exercise intensity | repetition-continuum based prescription (40~60% 1RM progressing to 70~85% 1RM), Rating of Perceived Exertion (RPE)-based prescription (RPE 3~5 on Category Ratio 10 scale progressing to RPE 6~8) |
| Exercise volume | 1~3 sets of 6~12 repetitions |
| Rest periods | within session (60~120s between sets and 3~5mins between exercises), between sessions (at least 48 h) |

#### Developing phase

Using topic guides, at least two rounds of focus groups comprised of older adults with possible sarcopenia will be conducted (Figure 6). There will be at least six older adults in each focus group during each round. The optimal time allotted for each focus group is 45 to 90 minutes, during which no more than twelve predetermined questions are discussed^70^. If face-to-face focus groups are prohibited by local epidemic policy, we will consider conducting online focus groups instead. The purpose of the first round of focus groups is to discuss the problems in the paper version of the initial intervention plan and modify it based on the suggestions of our participants to better fit the targeted population. Following this, we will create short videos based on the revised paper plan. The short videos will include two categories of short instructional videos for TikTok: health education and exercise (Figure 7). Then, the second round of focus groups will seek to modify the content of each short video further. If there are numerous issues with short videos, we will consider conducting a third round of focus groups to review all of the revised short videos and ensure that each segment is appropriate for our stakeholders and free of controversy.

**Figure 6.**
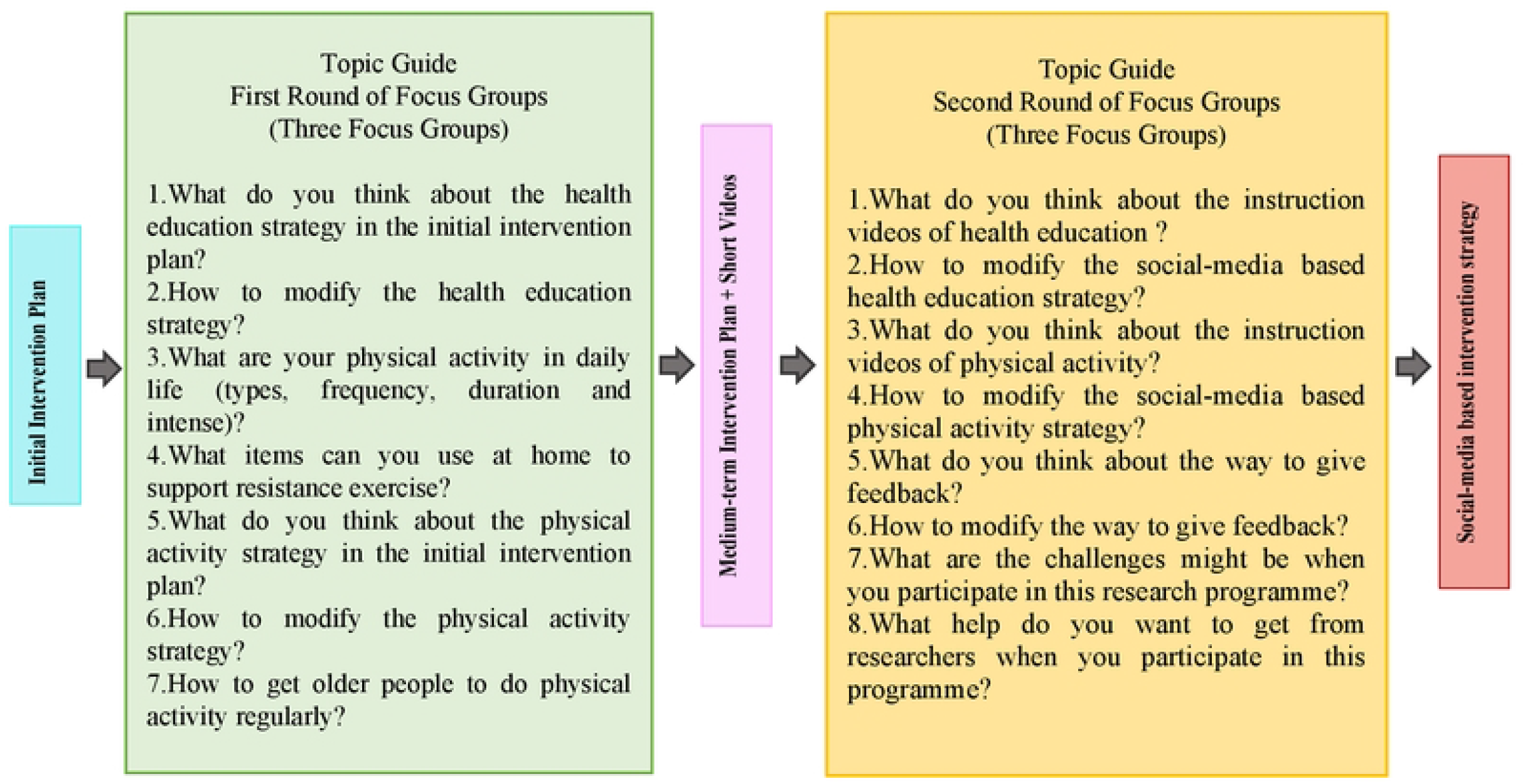
Topic Guide of Two Rounds of Focus Groups. NOTE: The topic guide described in this diagram is just a preliminary idea, and the specific details will be presented in the real practice stage.

**Figure 7.**
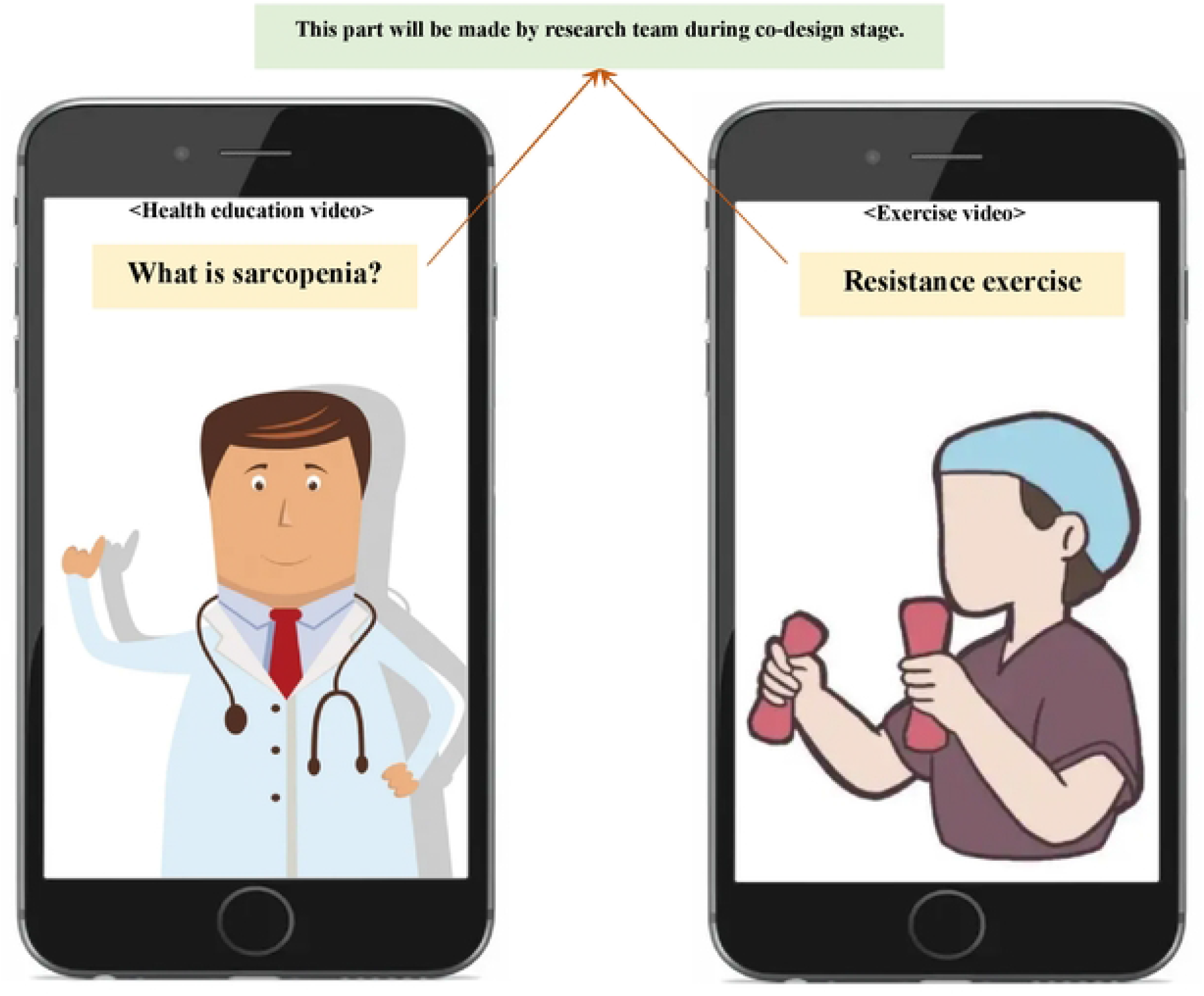
Instructional video examples for TikTok

#### Pre-testing phase

We will then obtain a fully theory-based conceptual model and a social-media based intervention strategy, which will be tested and modified in a small pre-feasibility study during three weeks. The primary observations and modifications pertain to the conceptual model, intervention strategy, and intervention process, such as the researchers’ guiding process and older adults’ learning process. Once these aspects satisfy the success criteria (Figure 8) for various stakeholders, we can proceed to the next stage, a feasibility study.

**Figure 8.**
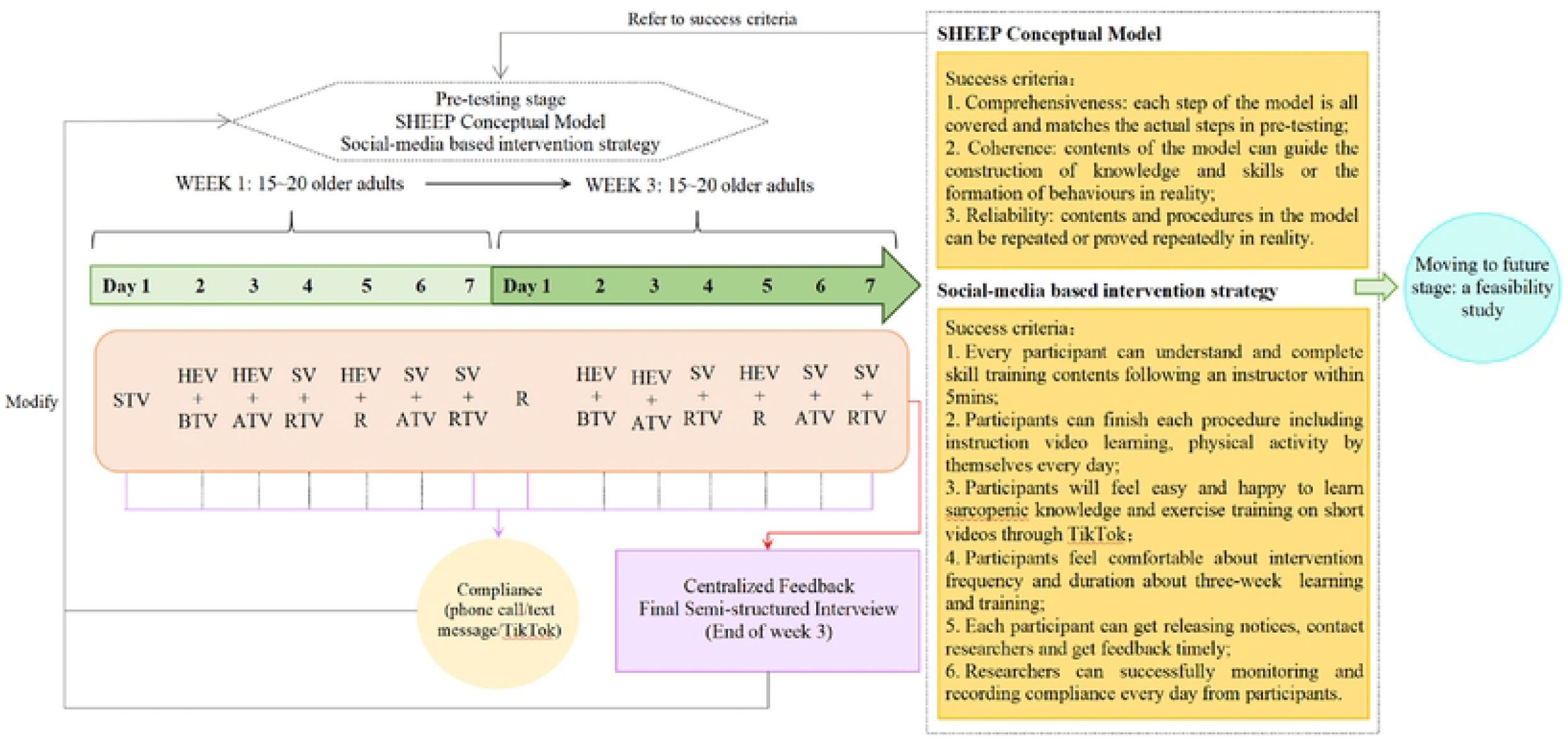
A brief diagram of the pre-testing procedure. Note: The intervention strategy in this diagram is just an example using for demonstration. STV, skill training video; HEY, health education video; BTV, balance training video; ATV, aerobic training video; RTV, resistance training video; SV, summary video; R, rest.

We will mainly collect two categories of data from participants, including intervention compliance and centralized feedback, in order to verify the appropriate contents and the correct dose of health education and exercise (in terms of frequency, intensity, and duration) can be achieved using a social-media-based approach.

1. Intervention compliance: Depending on their preference, participants can inform researchers of their completion of viewing instructional videos and physical activity via phone call or text message. To monitor participant compliance, the researcher will keep a diary for each participant using a dedicated mobile phone that is not a personal own. The researcher will purchase a new SIM card and dedicated phone number in China for this study.
2. Centralized feedback: This will be a semi-structured interview conducted face-to-face or over the phone in order to delve into the essence of problems and solicit suggestions for future revision. The semi-structured questions would be like: How do you feel throughout the entire pre-testing phase? What do you think about the social media-based health education in pre-testing phase? How to improve the social-media based health education? What do you think about the social-media based physical activity in pre-testing phase? How to improve the social-media based physical activity? In addition to health education and physical activity, what other aspects of this project do you believe require improvement? What do you believe the challenges will be if this programme is extended to a greater number of older individuals with possible muscle loss?

Moreover, researchers should document issues that arise during the guiding process, such as uploading videos to TikTok, distributing notices in TikTok groups, monitoring and recording compliance of behaviour change, and determining whether communication on TikTok is efficient.

#### Data collection and analysis

To promote the collection of high-quality data, the assessors will double-check the collected data, and a statistician will examine the range of values.

As for quantitative data analysis, descriptive statistics will be utilised to summarise the percentage of recruitment, baseline participant characteristics (e.g. age, gender, and educational background), attrition rate, compliance rate, etc. All statistical analyses will be conducted using SPSS Statistics 27.0 (IBM Corp., Armonk, NY, USA). P Values <0.05 will be interpreted as indicating significance.

Regarding the collection of qualitative data, all face-to-face recordings of group interviews in focus groups and final semi-structured interviews in the pre-testing phase will be audio recorded using an encrypted digital recorder. If face-to-face meetings are prohibited by local epidemic policy, the focus group will be conducted using Tencent Conference Software, which can encrypt meetings to protect participants’ privacy, and only audio of the group meeting will be captured via the cloud recording function. In addition, if participants prefer a telephone interview for the final semi-structured interview, the conversation will be recorded on an encrypted dictaphone. All encrypted recordings will be uploaded to a secure server at the University of Manchester and then deleted from the recording device and Tencent Conference cloud. Moreover, each interview will last between 30 minutes and one hour, and participants will only be asked to participate in one.

During the data analysis stage, qualitative data will be transcribed verbatim and potentially identifying information will be removed. The transcripts will be compared to the original recordings, corrected as needed, and anonymized. To become familiar with qualitative data, researchers must first read all transcripts. Themes will be developed based on the interview guide and research team discussion in order to identify the key characteristics of the qualitative data. Then, codes will be generated for each line of each transcript and categorised accordingly. Before finalising the major themes, each category must be reevaluated and scrutinised, and groups of major categories will be refined. Two researchers will perform and review the coding process to ensure a double check, followed by a discussion to ensure the validity of the data. To increase the transparency of the interpretation, the Chinese-to-English and English-to-Chinese translation of quotations will be performed in this study. QSR International’s NVivo 12 qualitative analysis software will be used to assist and facilitate the coding and analysis process.

## TRIAL MONITORING

The principal investigator will oversee participant recruitment, intervention development and pre-testing stage. In addition, one community leader in the cooperative community health centre will assume overall responsibility for participant identification, recruitment, and pre-testing. One team comprised of research professionals and academic experts from the United Kingdom and China will be responsible for ensuring the overall quality of research data. All principal researchers will hold a monthly online meeting to report on the project’s progress and discuss its problems and solutions. The team of professionals and experts will provide guidance and aid in dissemination. The principal investigator will review the risk register. Moreover, the study is subject to the audit and monitoring regime of the University of Manchester and the monitoring plan followed. A thorough risk assessment has been conducted, and potential patient, organisational, and study hazards, as well as their likelihood of occurrence and potential consequences, have been considered.

## RESEARCH ETHICS

This study has been approved by the University of Manchester Research Ethics Committee (Project ID: 15664), and permissions have already been granted by collaborators in relevant Chinese organisations.

## DATA MANAGEMENT

The principal investigator will maintain the confidentiality of study participants and adhere to the General Data Protection Regulation’s transparency requirements for health and care research. Data and all pertinent documentation will be confidentially and securely stored for a minimum of ten years following the conclusion of the study.

## DISCUSSION

In order to slow or even halt the progression of sarcopenia, it is crucial for healthcare professionals to design appropriate behaviour change interventions to help the targeted population increase awareness of sarcopenia prevention and form a habit of regular activity. Especially with the rapid development of information and communication technologies (ICTs) and the advent of pandemics in recent years, digital health is emerging as a key sector for delivering health services around the world, encompassing many facets such as e-health, mobile-health (m-health), medical informatics, and telemedicine^76,77^. The World Health Organization also announced a Global Strategy on Digital Health 2020-2024 with the mission to “improve health for everyone, everywhere by accelerating the adoption of appropriate digital health.^78^” Thus, digital health has the potential to be a paradigm shifter in terms of enhancing information, education, communication, health monitoring, diagnostics, and data management^79^. The SHEEP programme was conceived against such a backdrop, which has stronger competitiveness and considerable potential as it can meet the needs of the information age by providing targeted older populations with timely health education and exercise through social media that offer numerous advantages in terms of accessibility, time savings, cost savings, and continuity of care.

An integrated and reasonable behaviour change strategy considering stakeholders’ preferences may also be the key to the success of the SHEEP programme. To date, no intervention strategy has been developed to assess sarcopenia patients’ mastery of sarcopenic knowledge, investigate their attitudes towards sarcopenia, or monitor whether they continue to engage in healthy behaviours such as regular exercise after completing their studies. Hence, the SHEEP programme focuses on enhancing health education of sarcopenia prevention for the targeted population, with the goal of enhancing their knowledge of sarcopenia prevention and thereby strengthening their awareness and attitude towards sarcopenia prevention. This programme simultaneously encourages the target population to change their behaviour and develop a daily exercise routine. Furthermore, older population frequently experience age-related sensory (e.g., impaired vision and/or hearing) and cognitive declines (e.g., reduced memory and/or processing speed)^80-84^. Consequently, the single presentation of learning materials in oral or textual form and the overload of learning time, such as more than one hour at a time, may appear impersonal and less effective^82,85^. Therefore, the SHEEP programme attempts to compensate for the shortcomings of single text or audio materials or lengthy lessons with short videos, and to use co-design to solicit the opinions of the targeted population to ensure that the intervention strategy is acceptable to them and meets their needs.

This research has several limitations. First, as the proposed study focuses solely on intervention development, the sample size will be small; consequently, it may not be representative of the entire Chinese older population. Second, due to the short duration of the pre-testing phase, we will not draw any conclusions about the intervention effectiveness, which will be evaluated in a subsequent study. Third, the study findings are unique to the community setting and population in China and may not be generalised to other settings or countries. Nevertheless, this study will demonstrate whether the social-media based intervention content, frequency, duration, process are appropriate to young-older adults with possible sarcopenia.

## Data Availability

No datasets were generated or analysed during the current study. All relevant data from this study will be made available upon study completion.

